# Hemi-brain growth as a biomarker for whole brain growth

**DOI:** 10.1101/2025.09.28.25336850

**Authors:** Utkarsh S. Bajaj, Mingzhao Yu, Kelsey Tempelton, Srijit Mukherjee, Nichol Nunn, Abhaya V. Kulkarni, John Kestle, Vishal Monga, Steven J. Schiff

## Abstract

**Objective:** Accurate cerebrospinal fluid (CSF) and brain volume estimation are important components for evaluating hydrocephalus treatments, including shunts and endoscopic third ventriculostomy (ETV) procedures. While MRI-based segmentation typically provides precise measurements, metallic artifacts from implanted shunts in hydrocephalus patients can impede accurate volume determination. This study introduces a method for assessing brain growth in hydrocephalus patients using artifact-affected MRI scans and presents an efficient, automated artificial intelligence (AI)-based pipeline for hemi-brain segmentation and subsequent volume assessment.

**Methods:** The study consists of 75 patients participating in the Endoscopic versus Shunt Treatment of Hydrocephalus in Infants (ESTHI) trial. Pre- and post-operative T2 MRI scans were collected. Hemi-brain growth curves for the artifact-free hemisphere are proposed to assess postoperative brain growth from MRI with metallic shunt artifacts. An AI-based hemi-brain volume estimation pipeline was developed, consisting of a brain/CSF segmentation model and a hemi-brain mask generator. Segmentation labels, including left/right hemi-brain masks and brain/CSF segmentation maps were created. The AI pipeline was trained and validated using a manually segmented data subset. The volumes of left and right brain hemispheres after surgeries were calculated and analyzed.

**Results:** Postoperative hemisphere volume ratios approach the normal ratio and remained constant over time, confirming the feasibility for use of hemi-brain measurements as proxies for whole-brain volume assessment in the presence of metallic artifacts. Additionally, the AI-based pipeline demonstrated high accuracy in generating hemi-brain masks and segmenting brain/CSF, effectively automating the process of hemi-brain volume estimation.

**Conclusions:** The hemi-brain volume estimation of the unaffected hemisphere offers a feasible method for assessing brain growth over time. This process can be automated using a highly accurate AI pipeline, providing a valuable tool for monitoring brain growth in pediatric hydrocephalus patients with shunts.

## Introduction

Hydrocephalus is a neurological condition characterized by an excess of cerebrospinal fluid (CSF) within the brain. If the passage of CSF flow is impeded, an excess of fluid is produced, or the resorption process is inhibited, the fluid will accumulate within the closed space of the skull causing an increase in intracranial pressure and hydrocephalus.^1^ If left untreated, the excess fluid will can lead to permanent brain damage and, in the pediatric population, developmental impairments.^2–7^ Pediatric hydrocephalus affects 1-2 per 1,000 infants in the United States^8,9^, with higher rates in low- and middle-income countries^10^. There are two primary operative treatments for hydrocephalus: cerebrospinal fluid shunting and endoscopic third ventriculostomy (ETV), the latter often combined with choroid plexus cauterization (CPC) in infants.^11–13^ In standard practice, treatment efficacy has been assessed through clinical evaluation and ventricular size measurements such as the frontal-occipital horn ratio (FOHR)^14^. However, these methods inadequately capture the complex interplay between CSF dynamics, brain growth, and neurocognitive outcomes.^15,16^

Brain growth in children is under intense study as a potential adjunct in the management of hydrocephalus. In long-term follow-up of a cohort in an ongoing randomized controlled trial of shunt vs endoscopic treatment of postinfectious hydrocephalus in Africa ^41^, brain volume achieved at 2 and 5 years postoperatively, independent of operative procedure, was strongly associated with neurocognitive outcome.^17,18^

To assess image-based brain volume measurements, there has been considerable effort characterizing the normal curves for childhood brain growth, and the relationship between such growth and anthropomorphic measurements of body size.^19^ Although there are differences in lobar and sub-lobar ratios of growth during childhood, the total size of each hemisphere demonstrates remarkably equivalent volume growth in the normal child.^19^

Many shunt valves contain sufficient ferromagnetic metallic components to cause substantial artifact on MRI. If such artifacts render inaccurate the measurement of brain volume for one hemisphere, might hemi-brain volume measurement from the unaffected side opposite the shunt provide a reliable proxy for whole brain volume measurement? This question forms the hypothesis explored in this current study.

In order to improve the accuracy of brain/CSF volume measurement, various studies have used MRI segmentation techniques.^20–24^ These methods have significantly improved precise quantification of brain and CSF volumes, offering a more nuanced understanding of structural changes associated with hydrocephalus and its treatment.^11,15^ Studies have shown that metal-induced susceptibility artifacts can cause substantial signal loss, geometric distortion, and failure of fat suppression around shunt hardware^25^. The severity of artifacts increases with magnetic field strength, making 1.5T scanners preferable to 3T for imaging patients with metallic implants.^26^

While post-processing methods such as Virtual Brain Grafting^27^ (VBG) have shown promise in some applications, they have important limitations for hydrocephalus imaging due to ventricular and brain asymmetry, and significant anatomical distortions common in these patients. Machine learning-based inpainting techniques have been explored, but struggle with the complex, patient-specific nature of hydrocephalus-related brain changes.^28^ Despite ongoing research, there is no ideal solution for recovering artifact-corrupted regions in hydrocephalus imaging, as current methods such as digital tomosynthesis ^29^, magnetic resonance spectroscopy^30^, ultrashort echo time sequences,^31^ and k-space corrections ^32,33^ have significant limitations for this task.

To address these challenges, we propose the hemi-brain growth curve as a novel proxy for assessing brain growth by measuring the artifact-unaffected hemisphere. This approach is based on the fact that during normal childhood development, the volume of the left and right hemispheres tend to grow symmetrically,^19^ and we hypothesized that the hemispheric volume ratio following shunt surgery would stay approximately constant. In addition, we sought to develop an artificial intelligence (AI)-based pipeline enabling automatic, efficient and accurate hemi-brain volume estimation.

## Methods

This interim technical report presents findings from an ongoing randomized trial. Detailed information regarding the patients, both prior to and following randomization, will be provided in future publications but is not included in this report. The decision to publish this interim technical report stems from the potential importance and value to the field of our demonstration of a method to assess brain growth in the presence of metallic shunt artifact.

### MRI Subjects and Study Design

This study utilizes imaging data from the Endoscopic versus Shunt Treatment of Hydrocephalus in Infants (ESTHI) trial, a multicenter, prospective study designed to evaluate treatment outcomes in pediatric hydrocephalus (ClinicalTrials.gov registration NCT04177914). The trial was conducted at multiple tertiary care centers, each with Institutional Review Board (IRB) or Research Ethics Board (REB) approval. Informed consent was obtained from the legal guardians of all participants. The images studied in this present analysis were from children less than 2 years of age receiving first time treatment for hydrocephalus.

### Ground Truth Labels and Hemi Brain Mask

Ground truth labels, i.e. brain/CSF segmentation label masks and unaffected hemi-brain label masks for each MRI scan, were manually created by a neurosurgical resident and subsequently verified by a senior neurosurgeon to ensure accuracy. Manual segmentation was performed using ITK-SNAP software, which facilitated precise delineation of anatomical structures necessary for brain volume assessment.^33^

### Constant Hemisphere Volume Ratios of Postoperative Hydrocephalus Patients

Normal brain structures during development do not exhibit perfect symmetry.^35^ However, the ratio of left to right brain volumes tends to remain constant for normal male and female children, as evidenced by data reanalyzed (Figure 1).^19^ We therefore hypothesized that the hemi-brain left to right volume ratio of each postoperative hydrocephalus patient is also approximately constant as the brain grows. If so, individual hemispheres may contain sufficient information for monitoring brain growth, serving as a proxy for whole brain growth.

**Figure 1.**
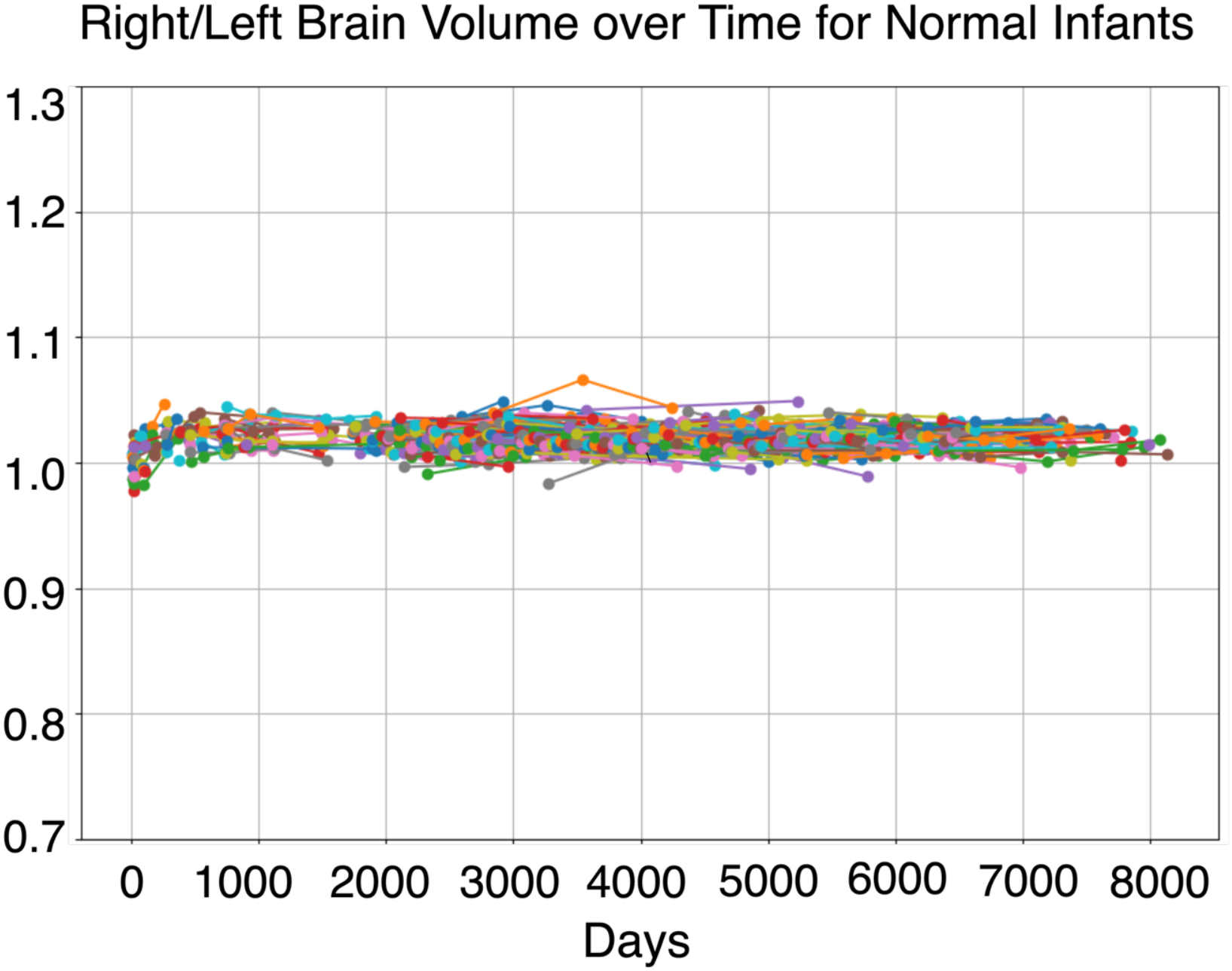
Left/Right Brain Volume Ratio Over Time. This figure illustrates the left-to-right brain volume ratio over time for normal brains, showing that the ratio remains consistent.^37^ This stability demonstrates the feasibility that hemisphere volume tracking can reliably estimate whole brain growth.

We designed statistical tests to investigate the hypothesis that the distribution of the hemi-brain volume ratios across the patients is maintained at postoperative time-points. By systematically dividing the data into distinct time ranges—preoperative (Days after intervention = 0), early postoperative (0 < Days after intervention ≤ 180 days), and late postoperative (Days after intervention > 180 days)—we ensured a structured approach to analyzing the trends in hemi-brain volume ratios. For all the patients reported in this study, the operative side was the right, and the non-operated side was the left side of the brain. This step-by-step temporal division provided a clear basis for statistical analyses, including Mann-Whitney U-Test, which demonstrated that while the postoperative ratios remained stable over time, the preoperative ratio exhibited statistically significant changes from the postoperative ratios.

### Hemi-brain Growth Curve and an AI-based Pipeline

Due to the statistically verified constant ratio between hemispheres in postoperative hydrocephalus patients, we propose the Hemi-brain Growth Curve for those patients without measurable whole brain volumes due to metallic shunt artifacts.

An AI based brain volume estimation pipeline for post-operative MRI with shunt artifact was constructed. The pipeline consists of three procedures: 1) obtain brain/CSF segmentation maps 2) obtain hemi-brain masks of the artifact-unaffected side 3) calculate the brain and CSF volume of the hemi-brain using the aforementioned segmentation maps and voxel information, as visually represented in (Figure 2).

**Figure 2.**
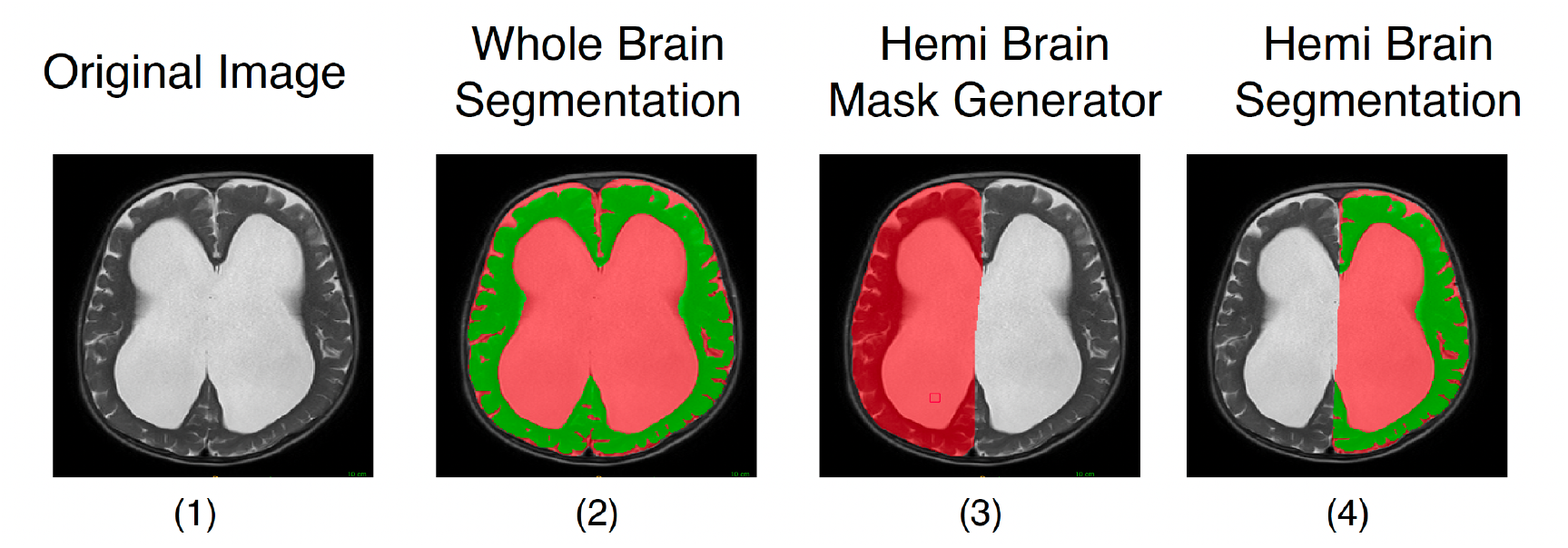
The figure illustrates the process of calculating hemi-brain volume, showing (1) the original image, (2) whole-brain segmentation, (3) hemi-brain mask, and (4) hemi-brain segmentation derived by applying the hemi-brain masks to the whole-brain segmentation.

To automate procedures 1 and 2, a deep learning (DL)-based whole brain MRI segmentation model and a DL-based hemi-brain mask generator were developed and trained respectively. The model adapts a 2D fully convolutional DenseNets architecture with Encoder Decoder structure similar to 2D U-Net.^36,37^ Two such neural network architectures are built, to automatically segment T2-weighted brain image scans into three classes: brain tissue, cerebrospinal fluid (CSF), and background, and a network to generate hemi-brain masks.

To calculate the brain and CSF volume, the segmentation area of each axial slice was multiplied by the slice thickness, and these values were then summed across all slices. Appendix Figure 1 contains more details about the model. The model’s encoder-decoder structure maps each 2D image into important features, which are then expanded to a segmentation map with the same resolution as the original image. The segmentation image contains the probability of each pixel being brain tissue, CSF, or background. The loss function consists of Dice similarity loss^38^ for the segmentation output and Total Variation loss^39^ of decoder features, including the segmentation output. This function is minimized to generate segmentation masks on unseen data. The AI model was trained using 80% of the ESTHI dataset, with rigorous validation on an independent 10%, and 10% as a hold-out test set using one NVIDIA Titan X GPU. Further details about the deep learning method and training procedure are described in the Appendix.

## Results

Preoperative MRI scans of pediatric patients (Figure 3a–d) illustrate typical brain region visualization prior to surgical intervention. However, the presence of metallic artifacts— particularly from unilateral shunt valves—substantially degraded image quality in affected regions (Figure 3e–h), rendering up to 30% of the brain volume non-diagnostic. It is noteworthy that any artifacts observed did not affect the contralateral hemisphere.

**Figure 3.**
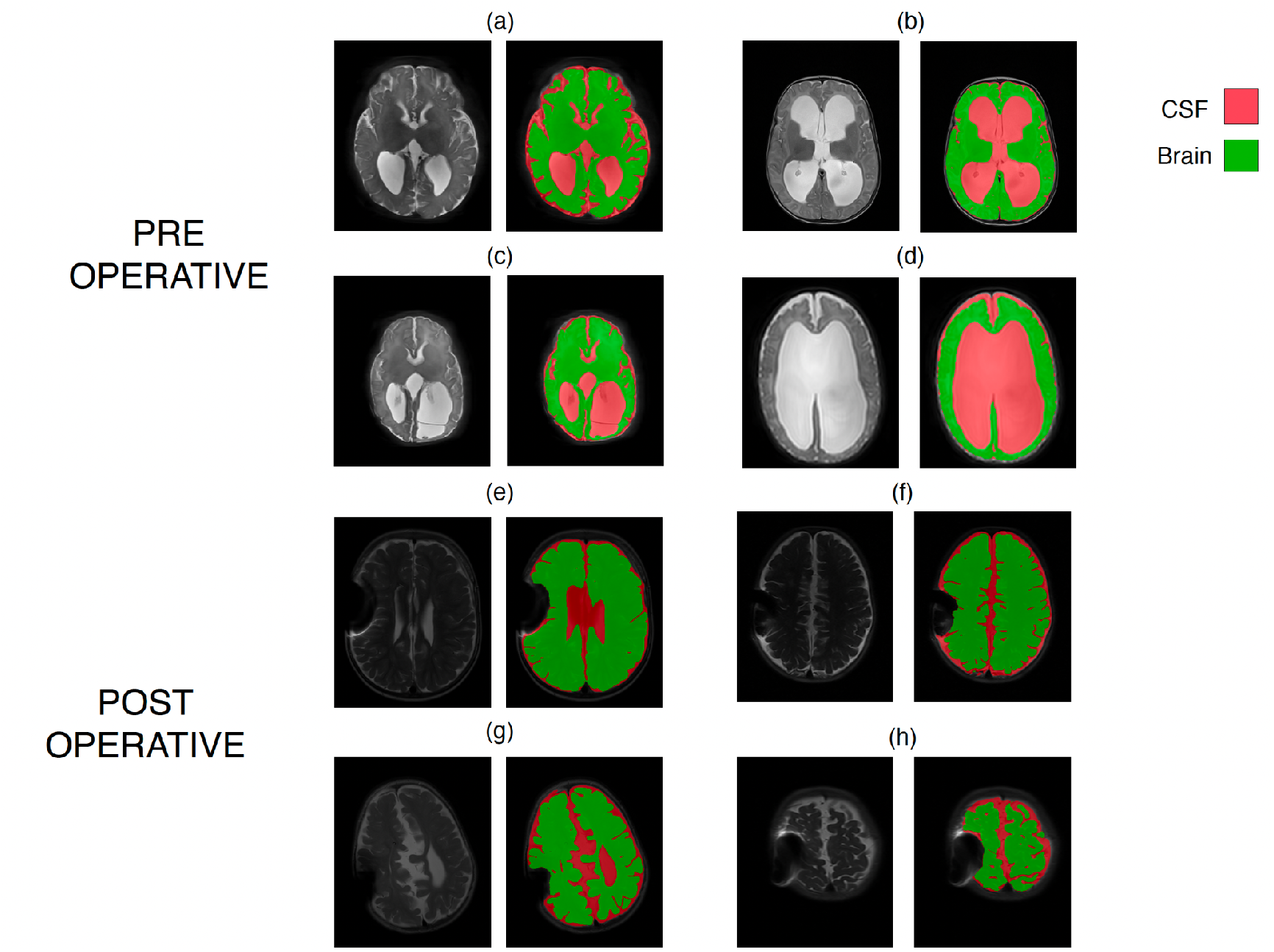
MRI Scans from the ESTHI Trial. This figure illustrates selected data from the ESTHI trial, comprising 361 sets of MRI scans from 75 patients. (a), (b), (c), (d) represent 4 preoperative images and their segmentation into brain and CSF. Each scan includes approximately 30 T2-weighted axial scan slices, each with a resolution of 500 x 500 pixels. Manual labeling was performed, assigning each pixel within the brain to either cerebrospinal fluid (CSF) or brain tissue. (e), (f), (g), (h) represents the impact of metal susceptibility artifacts on MRI Scans from implanted shunt hardware. Approximately 50% of subjects treated with shunts were affected by these artifacts, which prevented the accurate measurement of whole brain volume and growth for these individuals.

### Visual Consistency of Hemisphere Ratios

Analysis of MRI data without metallic artifacts from the ESTHI trial revealed that the volume ratio between the non-operated and operated hemispheres stabilized over time following surgery (Figure 4). The consistency of these ratios over time suggests that hemi-brain measurements can serve as a proxy for whole-brain volume assessments.

**Figure 4.**
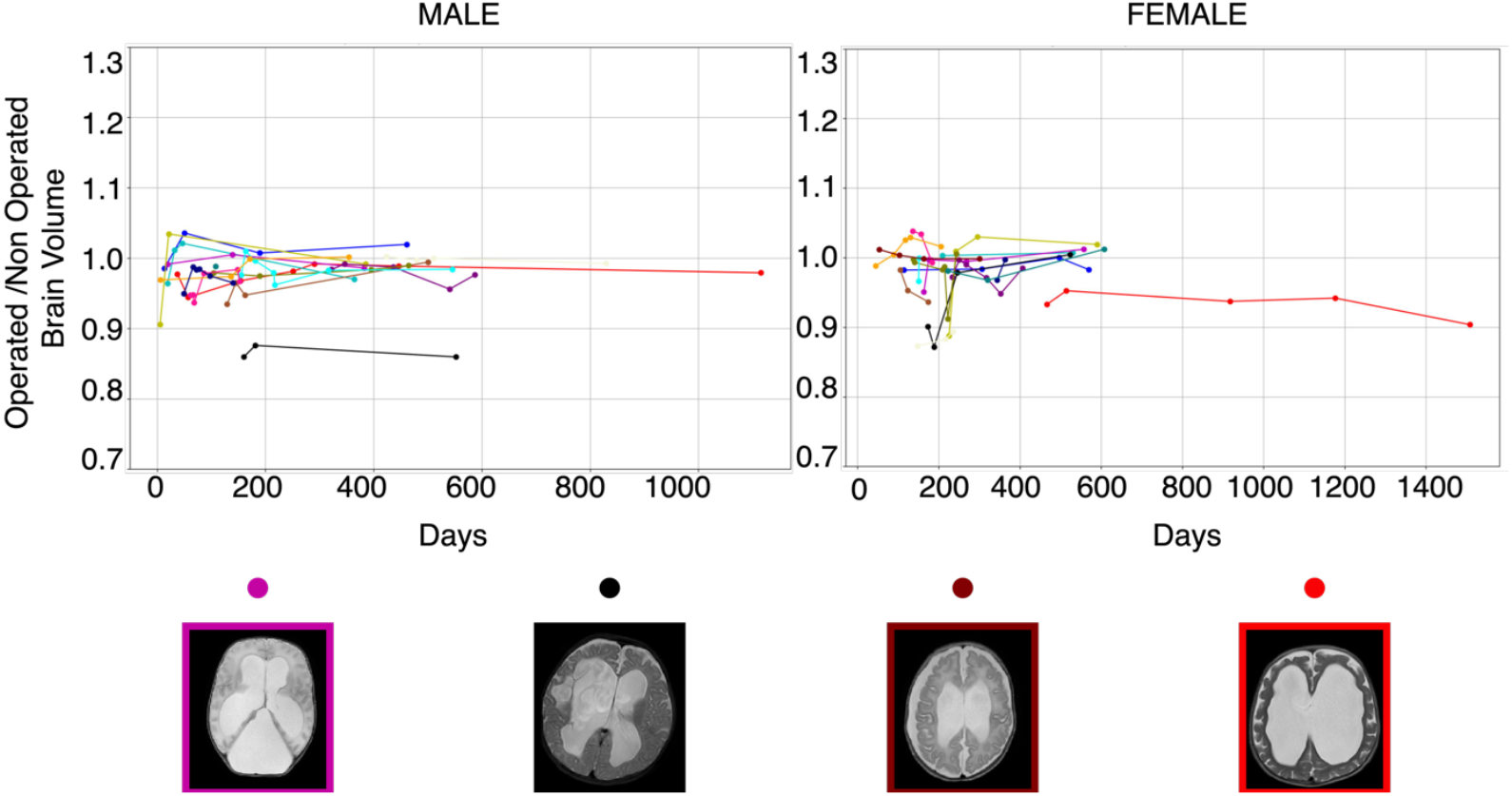
Hemi-Brain Analysis for Estimating Brain Growth. This figure illustrates examples using hemi-brain volume analysis to estimate brain growth over time. Some subjects demonstrated perturbations in operated/non-operated volumes early after surgery, but these volume ratios stabilized over time. Such rapid perturbations may have been related to the effects of pressure changes and tissue edema before and after surgery. Note the examples of symmetric vs non-symmetric hemi-brain structure shown in the examples. The substantial asymmetry in hemi-brain ratios in the black trace in the left panel is readily seen in the example slice, but the more subtle asymmetry in the red trace in the right panel is less obvious to visual inspection prior to segmentation and quantification.

### Statistical Analysis of Consistency of Hemisphere Ratios

Statistical analyses (with a significance level of 95%) were conducted to validate the stability of hemisphere volume ratios over time. Figure 5 illustrates the “Operated by Non-Operated Brain Volume Ratio” across pre-operative, early post-operative, and late postoperative phases as represented in Table 1. Statistical analysis shows no significant difference between early post-operative (Days after intervention = 0–180 days) and late postoperative (Days after intervention >180 days) ratios (U = 1326.0, p = 0.4858). However, a Mann-Whitney U-Test comparing pre-operative (Days after intervention = 0 day) and post-operative (>0 days) ratios reveals a statistically significant difference (U = 1879.5, p = 0.0288), indicating that preoperative ratios can adjust significantly early post-surgery, prior to stabilizing.

**Table 1:**
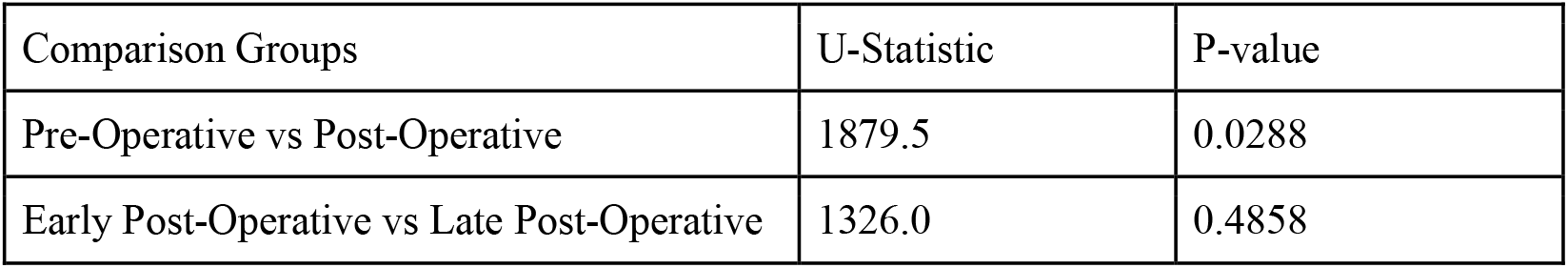
Statistical comparison of Operated to Non-Operated Brain Volume Ratios over time. AI.

| Comparison Groups | U-Statistic | P-value |
| --- | --- | --- |
| Pre-Operative vs Post-Operative | 1879.5 | 0.0288 |
| Early Post-Operative vs Late Post-Operative | 1326.0 | 0.4858 |

**Figure 5.**
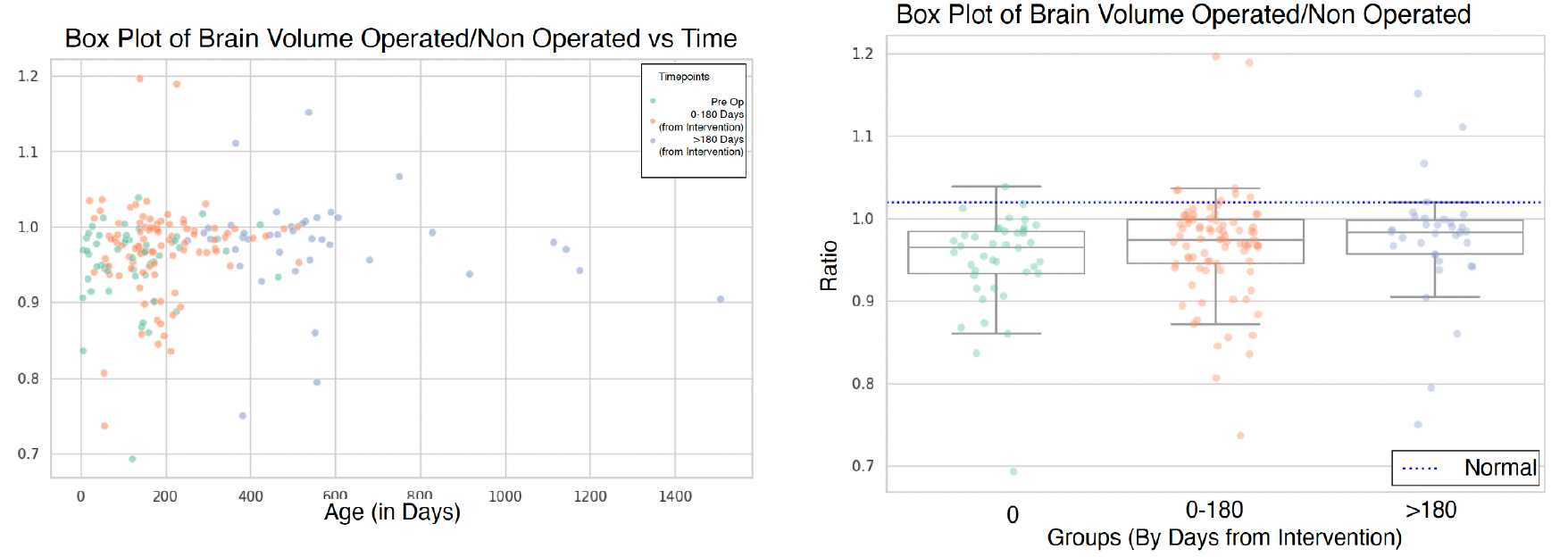
Operated vs Non-Operated Brain Volume Ratio over time. The figure shows the ratio of operated vs non-operated brain volume ratio across three time ranges: pre-operative, early post-operative(0 < Days from intervention < 180), and late postoperative (Days from intervention > 180). The post-operative ratio tends to normalize over time, approaching a more symmetric ratio near 1.0 (Mann-Whitney UU = 1879.5, p = 0.0288). The median right/left median ratio for normal children is shown as a dashed line.^21^

### Model Performance

The AI-based pipeline demonstrated accurate performance in segmenting brain structures, as evidenced by quantifying the overlap of pixels using the Dice similarity test. Specifically, the model achieved Dice scores of 94.7% for brain tissue and 94.2% for cerebrospinal fluid (CSF), indicating a high level of accuracy in segmentation tasks. The use of a 2D fully convolutional DenseNet architecture (Figure: Appendix Figure 1) facilitated accurate automated segmentation, which is crucial for reliable volume estimations in clinical settings. The AI model for hemi brain mask generation demonstrates not only robustness to angular variations, as seen in Figure 6, but also consistent accuracy across the full brain volume, as illustrated in Figure 7, achieving a Dice score over 94% with only minimal manual adjustments applied to enhance precision, particularly in challenging areas.

**Figure 6.**
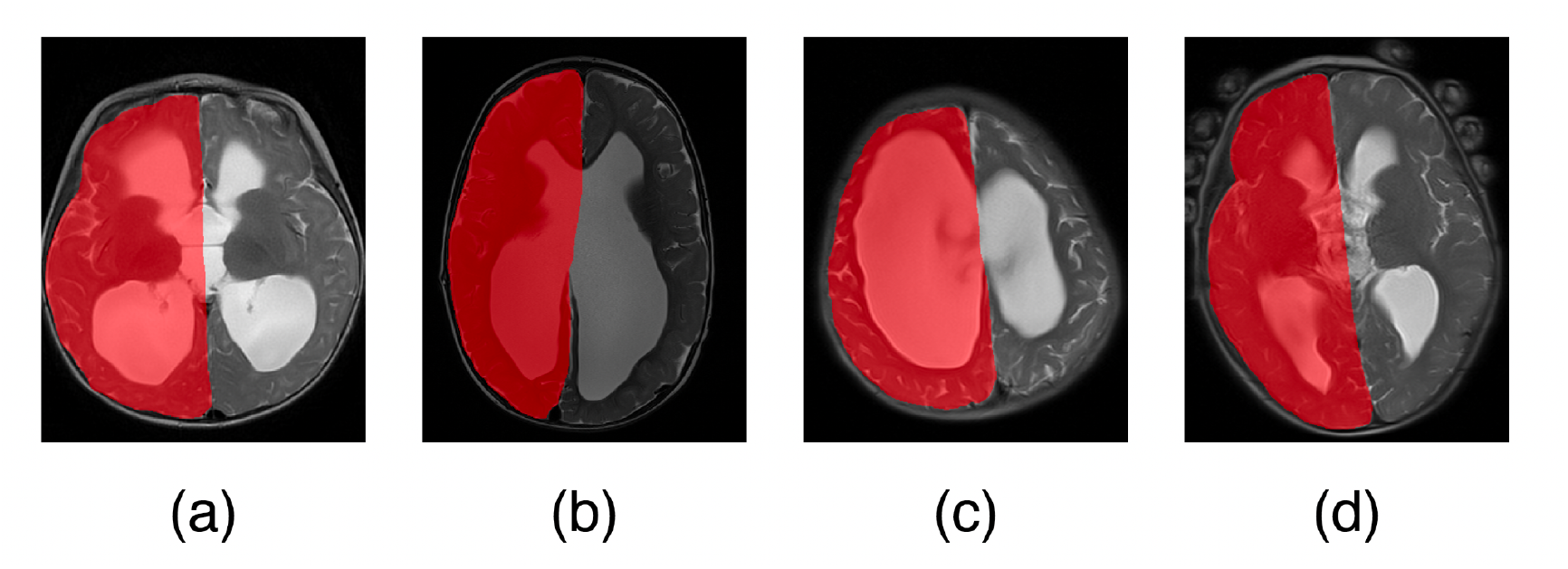
Hemi Brain Masks Across Varying Scan Angles. This figure visually demonstrates the robustness and accuracy of the hemi brain mask generator, even when applied to scans acquired at different angles. The generated masks consistently align with the anatomical structures, highlighting the model’s adaptability to varying orientations.

**Figure 7.**
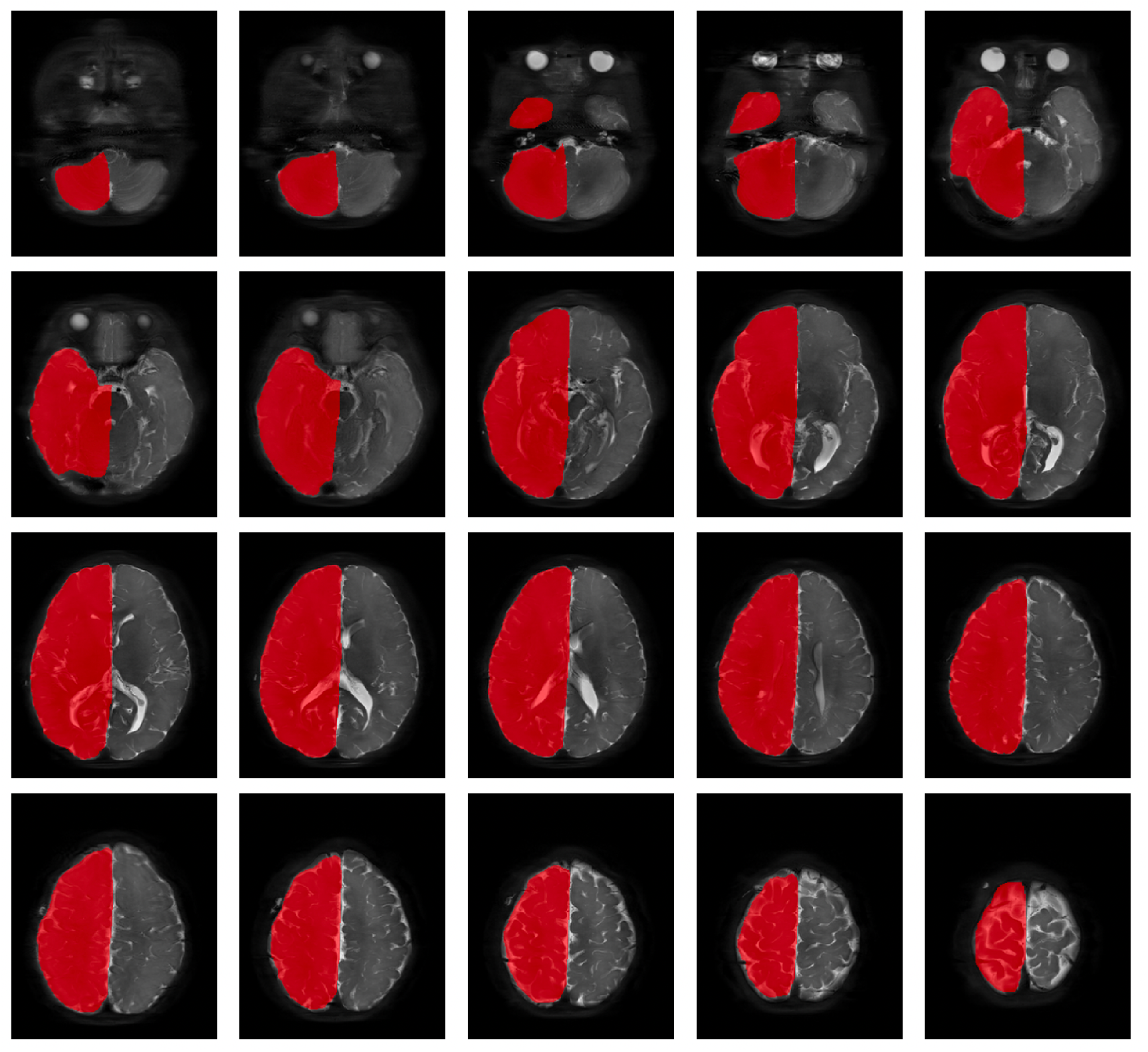
Hemi Brain Mask for Full Scan Set of One Subject. This figure demonstrates the performance of the hemi-mask generator across all axial slices from the top to the bottom of the brain for a single subject. The masks show consistency and accuracy in delineating the hemispheres throughout the entire brain volume. While the generator performed effectively in most regions, slight manual adjustments were applied to enhance precision, particularly in challenging areas.

### Hemi- and Whole-Brain Growth Curves

In patients without hemispheric metallic artifact, including both ETV/CPC and shunt valves that did not generate substantial artifact on MRI, the patterns of growth were similar between whole- and hemi-brain generated volumes (Figure 8).

**Figure 8.**
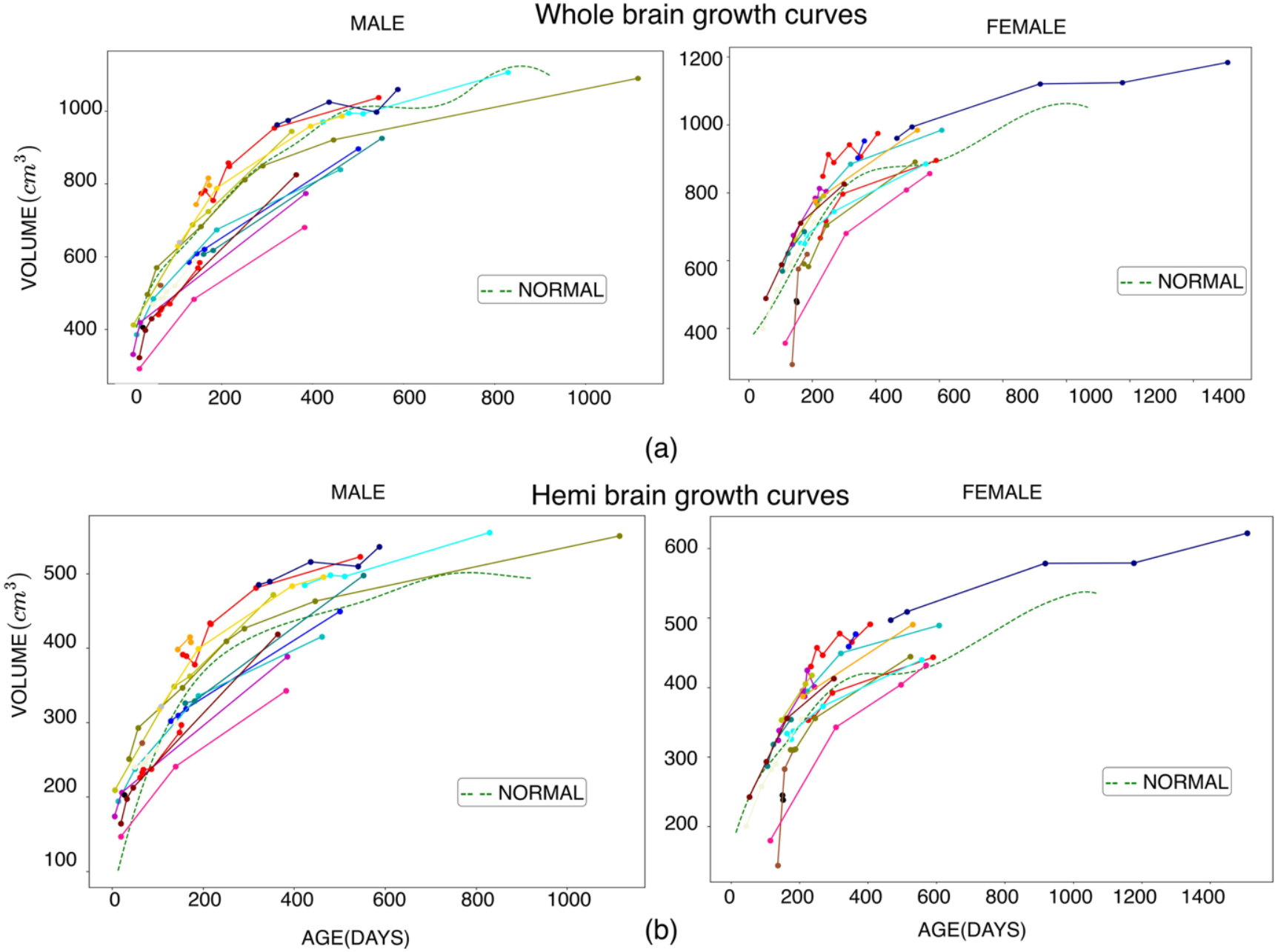
Whole- and Hemi-Brain Volume Growth Curves by Sex. (a) Whole-brain volume growth curves for male and female subjects treated with Endoscopic Third Ventriculostomy (ETV) or shunt, in cases without metallic artifacts. The analysis includes a comparison with normal brain volumes (dotted line). (b) Illustrates the growth trajectories of hemi-brain volumes for male and female subjects over time.

## Discussion

Our findings demonstrate that hemi-brain volume measurements can serve as a proxy for whole-brain volume measurements. This is valuable when a substantial shunt valve metallic artifact obscures portions of the brain on postoperative MRI. This strategy is possible because such artifacts on MRI were not found to affect the contralateral hemisphere.

### Significance of Hemisphere Ratio Stability

The early perturbation effects of surgery, with distortions of brain and fluid, are readily seen in the early changes in the operated/non-operated volume rations in Figure 4; nevertheless, these ratios then appeared to stabilize in patients over the follow-up time period.

Unexpectedly, we observed a normalization of the operated/non-operated side volume ratios during follow-up, seeming to approach a more equal ratio (approaching 1 in Figure 5), similarly to normal children’s hemispheric ratios ^19^. Although statistically significant, we do not understand what underlying regulatory mechanism might be active in achieving more symmetrical hemispheric brain growth in these children postoperatively.

### AI Pipeline Efficacy and Clinical Integration

Manual segmentation of brain structures is time-consuming and slow. The AI-based pipeline developed for this study proved highly effective, achieving a Dice similarity coefficient greater than 0.93 for segmentation accuracy on hold-out dataset testing. This level of precision underscores the potential of AI models to streamline and automate hemi-brain volume estimation for clinical use.

The hemi-brain volume pipeline was robust in the setting of inconsistent MRI scan orientation. Inconsistent head position is more common in pediatric brain scans. And with the increased interest in using low-field MRI for clinical evaluation of patients, robustness to lower scan resolution and contrast will be further helpful.^40^

### Broader Clinical Implications

The use of brain volume measurement in the management of hydrocephalic patients is currently under intense study.^17–18,41–44^ The ability to bypass the limitations of metallic artifacts in MRI imaging expands the inclusivity of volumetric brain growth analysis in hydrocephalus management to patients following shunt insertion.

### Limitations and Future Directions

While our study provides a promising approach to overcoming metallic artifact technical challenges, there are several limitations. The assumption of stable postoperative hemisphere ratios may not universally apply, particularly in cases with complex or asymmetric brain development. In addition, some occasional manual segmentation intervention remains necessary for all segmentation algorithms of pathological scans when the individual patterns are unique and were not well represented in the training data set. Additionally, while our AI pipeline demonstrated high accuracy, further validation on larger and more heterogeneous datasets is necessary to confirm its scalability. Future studies should also investigate the integration of this approach with complementary imaging modalities, such as ultrashort echo time sequences or advanced post-processing techniques, which could further mitigate the impact of metallic artifacts and enhance volumetric assessments.

### Conclusions

This study establishes hemi-brain volume measurements as a feasible proxy for whole-brain growth in pediatric hydrocephalus patients in the presence of metallic MRI artifacts. The development and validation of an AI-based segmentation pipeline provides a useful tool for automating high-accuracy hemi-brain volume assessments.

## Data Availability

All data produced in the present study are available upon reasonable request to the authors.

## Acknowledgements

Supported by NIH Grant 1U01NS107486.

Appendix

### Brain and CSF Segmentation

#### Architecture

We developed a fully convolutional Dense U-Net that leverages DenseNet-style^1^ skip-connected blocks within an encoder–decoder framework to segment brain tissue, cerebrospinal fluid (CSF), and background in medical images. Densely connected layers promote feature reuse and stable gradients, crucial for capturing the subtle intensity differences of CSF. Training minimizes a composite loss that sums the Dice coefficient— which drives accurate overlap between predicted and true masks—with a selectively applied total-variation (TV) term. TV is computed at every decoder up-sampling stage on feature maps masked to emphasize CSF-affected regions, penalizing abrupt pixel-to-pixel changes and yielding smoother, biologically plausible boundaries. This joint Dice + TV objective lets the network balance precision and regularity, ultimately producing cleaner, more reliable brain-scan segmentations.

#### Dice Coefficient

The Dice coefficient is used to measure the overlap between the predicted segmentation and the ground truth (true segmentation) for different parts of the image. It is defined mathematically as:

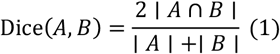

Where:

- *A* is the set of predicted pixels.
- *B* is the set of ground truth pixels.
- | A ∩ B | is the number of pixels in both sets (the intersection).
- |A| and |B| are the total number of pixels in the predicted and true segmentation, respectively.

The Dice loss can be formulated as:

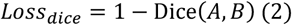

#### Total Variation (TV) Regularization

Total Variation (TV) Regularization helps to smooth the output by reducing noise and irregularities. It is calculated based on the differences between neighboring pixels. For a given feature map, the TV loss is defined as:

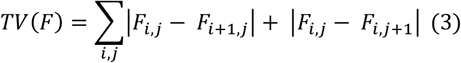

- where *F*_*i*,*j*_ represents the pixel value at position ***(i, j)***
- The summation runs over all pixels in the feature map, accounting for differences in both horizontal and vertical directions.

In the context of the model, TV is applied selectively in regions influenced by CSF using a binary region mask:

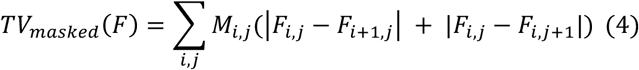

#### Final Loss Function

The final loss function combines both the Dice loss and the TV loss. It can be expressed mathematically as:

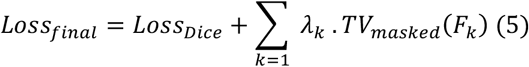

- where *λ*_*k*_ is a hyperparameter that controls the weight of the TV regularization in the overall loss for each *F*_*k*_ from the encoding block, allowing for a trade-off between segmentation accuracy and smoothness of the output.

In summary, the method minimizes the combined loss function, which is a sum of the Dice loss and the masked Total Variation loss, during training. This process enables the model to achieve a balance between accurately segmenting the regions of interest, such as CSF, and ensuring that the segmentation boundaries are smooth and biologically meaningful, thereby enhancing the quality and reliability of the medical image analysis.

**Appendix Figure 1:**
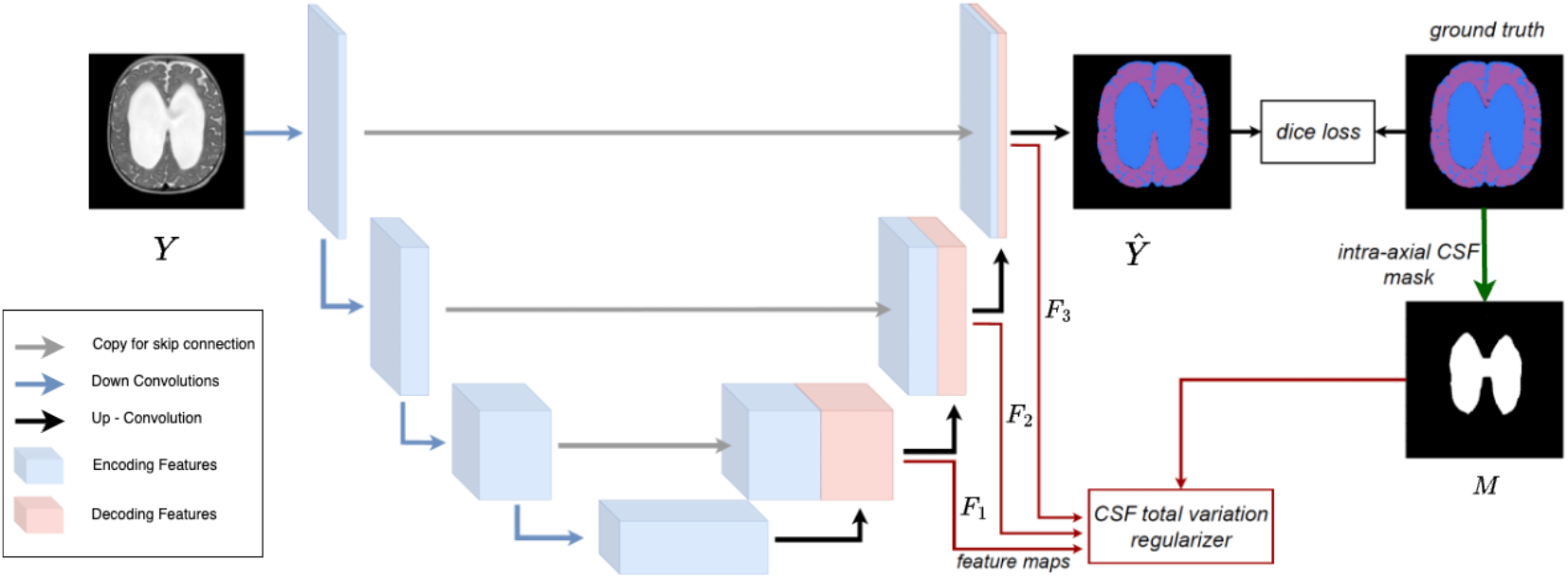
Regularization of Dense3 U-Net Outputs for CSF Region. This figure illustrates the regularization process applied to the Dense3 U-Net’s intermediate and final outputs. The regularization aims to reduce the total variation (TV) within the cerebrospinal fluid (CSF) region, resulting in smoother and more consistent predictions for this area of the brain.

